# Shorter leukocyte telomere length is associated with distinct CSF biomarker dynamics across early AD stages in at-risk individuals

**DOI:** 10.1101/2024.12.03.24318248

**Authors:** Blanca Rodríguez-Fernández, Armand González-Escalante, Patricia Genius, Tavia Evans, Paula Ortiz-Romero, Carolina Minguillón, Gwendlyn Kollmorgen, Nicholas A. Ashton, Henrik Zetterberg, Kaj Blennow, Juan Domingo Gispert, Arcadi Navarro, Marc Suárez-Calvet, Aleix Sala-Vila, Marta Crous-Bou, Natàlia Vilor-Tejedor, ALFA study

## Abstract

**INTRODUCTION:** Telomere length (TL) is a hallmark of biological aging. Shorter TL has been linked to an increased risk of Alzheimer’s disease (AD), but its role in AD pathophysiology remains unclear. This study investigates the relationship between TL, longitudinal cerebrospinal fluid (CSF) AD biomarkers, and brain structure in cognitively unimpaired (CU) individuals at risk for AD.

**METHODS:** We analyzed data from 346 middle-aged CU ALFA+ participants, measuring leukocyte TL (LTL) by qPCR. AD-related CSF biomarkers were measured at baseline and after 3 years. Stratified analyses by *APOE*-e4 and amyloid-tau (AT) status were conducted.

**RESULTS:** Shorter LTL was associated with higher astrocytic reactivity and synaptic dysfunction biomarkers, as well as thicker cortex in AD-vulnerable regions. Astrocytic biomarkers mediated the LTL-cortical thickness association. In *APOE*-e4 carriers and AT-positive individuals, shorter LTL linked to higher p-tau181 and neurodegeneration markers.

**CONCLUSION:** These findings highlight telomere shortening as a potential contributor of early AD-related progression.

**Highlights:**

- Shorter leukocyte telomere length (LTL) was associated with higher levels of cerebrospinal fluid (CSF) GFAP, CSF S100B and CSF α-synuclein, independently of amyloid and tau pathology.
- Shorter LTL was associated with higher baseline CSF NfL and t-tau levels in the A+T- and A+T+ groups, respectively.
- LTL association with brain structure was partially mediated by CSF biomarkers of astrocytic reactivity.

**Research in context:**

1. **Systematic review:** Literature review was performed using traditional sources (e.g., PubMed). While the association between leukocyte telomere length (LTL) shortening and increased sporadic AD risk is well-documented, its role in AD pathogenesis remains unclear. These findings have been appropriately referenced.
2. **Interpretation:** In cognitively unimpaired adults at higher risk for AD, shorter LTL was associated with high AD-related cerebrospinal fluid (CSF) biomarkers, including p-tau181 and biomarkers of neurodegeneration, synaptic dysfunction, glial reactivity, and inflammation. These associations were either more pronounce or exclusively observed in *APOE*-e4 carriers and individuals with early AD pathology (measured by CSF Aβ42/40 and p-tau181). Furthermore, increased astrocytic reactivity mediated the relationship between LTL and brain structure integrity.
3. **Future directions:** Further research is needed to understand the role of peripheral aging in AD pathology. Investigating how peripheral immune aging influences brain homeostasis and AD progression could help identify early targets of neuroinflammation and neurodegeneration.

## 1. Background

Biological age is a more accurate indicator of an individual’s physiological and functional status than chronological age [1,2]. Since aging is the major risk factor for developing neurodegenerative diseases, including dementia due to Alzheimer’s disease (AD) [3], significant efforts have focused on understanding how hallmarks of biological aging contribute to neuropathological processes [4–6].

Telomere length (TL) is a well-known hallmark of cellular aging [7]. Telomeres, composed of repetitive DNA sequences (TTAGGG) at the ends of linear chromosomes [8], are essential for genomic stability during cellular division [9,10]. Telomeres shorten with each cell division, a process accelerated by factors such as oxidative stress or environmental exposures [11]. Telomere shortening may induce DNA damage and cellular senescence, contributing to the aging process [12] along with other hallmarks such as epigenetic alterations [13].

Leukocyte telomere length (LTL) serves as a surrogate for TL in other tissues. While TL varies across tissues, it shows moderate positive inter-tissue correlations and an inverse relationship with age, the primary determinant of TL across tissues. Notably, although tissues with higher cell division rates, such as leukocytes, exhibit shorter TL, age-related TL shortening is also observed in tissues with a high proportion of post-mitotic cells, such as the hippocampus [14].

Shorter LTL has been linked to an elevated risk of mortality and age-related diseases, including all-cause dementia and dementia due to AD [15,16]. Cross-sectional and longitudinal studies indicate that shorter LTL, or greater LTL attrition, is associated with poorer cognitive performance in healthy young and middle-aged adults [17,18]. Furthermore, longer LTL is associated with larger grey matter (including the hippocampus) and white matter volumes, and cortical thickness [19].

Shorter LTL has been observed among AD patients when compared to controls [20,21]. Additionally, shorter LTL was found to predict a 25-year risk of AD incidence in *APOE*-e4 carriers in the prospective Betula project [22]. However, faster LTL attrition was only marginally associated with progression from normal cognition to mild cognitive impairment (MCI) and AD in the AD Neuroimaging Initiative (ADNI) cohort [21]. Moreover, no significant associations were found between LTL and cerebrospinal fluid (CSF) amyloid-β (Aβ) 42 or total tau (t-tau). Paradoxically, longer LTL was associated with cognitive decline in individuals with lower baseline CSF Aβ42 and higher CSF t-tau, and greater medial temporal lobe thinning among biomarker-negative ADNI participants [23].

AD is conceptualized as a biological and clinical *continuum*, driven by multiple interconnected and dynamic trajectories of pathophysiological changes, biomarker alterations, and cognitive symptoms, which collectively facilitate the transition from a prolonged asymptomatic stage to cognitive impairment [24–29]. Interactions between AD-related pathology, genetic factors, environmental influences, and hallmarks of aging are hypothesized to contribute to disease progression along this *continuum* [7,30,31].

Given these complexities and the paradoxical findings described above, there is a critical need to integrate longitudinal, multimodal biomarkers - beyond Aβ and tau-as well as AD-related neuroimaging endophenotypes to better elucidate the role of LTL in AD pathophysiology. Furthermore, focusing on the earliest stages of the AD *continuum* may help clarify the biological pathways through which aging contributes to exacerbate AD pathology and progression to dementia, while also identifying potential resistance and resilience mechanisms that support successful cognitive aging [32].

In the present study, we explored the association between LTL and both baseline levels and 3-year changes in AD-related CSF biomarkers, including Aβ and tau pathology, neurodegeneration, astrocytic and microglial reactivity, and inflammation. We also examined LTL associations with baseline cortical thickness in brain regions associated with aging and AD-related atrophy, as measured by magnetic resonance imaging (MRI). Finally, we evaluated whether *APOE*-e4 carriership and amyloid-tau (AT) pathology influenced these associations. This approach aims to disentangle transient signals and confounding factors from the underlying biological mechanisms linking shorter LTL to an increased risk of late-life dementia.

## 2. Methods

### 2.1. Study population

The present study was performed in the ALFA+ cohort. The ALFA+ study includes a subset of 450 participants from the Alzheimer’s and Families (ALFA) study [33] who were invited to participate based on their specific AD risk profile. Inclusion criteria considered participants’ AD parental history, *APOE e4* status, verbal episodic memory score and Cardiovascular Risk Factors, Aging, and Incidence of Dementia (CAIDE) score. A comprehensive characterization was performed in ALFA+ participants, including demographic characteristics, anthropometric measurements, a lumbar puncture for the measurement of CSF biomarkers and imaging (i.e., MRI and positron emission tomography (PET)) biomarker acquisition. ALFA+ inclusion criteria were: (1) individuals who had previously participated in the ALFA study; (2) age between 45 and 65 years at the moment of inclusion in ALFA; and (3) long-term commitment to the study: inclusion and follow-up visits and agreement to undergo all tests and study procedures (MRI, PET and lumbar puncture). ALFA+ exclusion criteria were: (1) cognitive impairment (Clinical Dementia Rating (CDR) >0, Mini-Mental State Examination (MMSE) <27 or semantic fluency <12); (2) any systemic illness or unstable medical condition that could lead to difficulty complying with the protocol; (3) any contraindication to any test or procedure; and (4) a family history of monogenic AD [34].

The ALFA+ study (ALFA-FPM-0311) was approved by the independent ethics committee ‘Parc de Salut Mar’, Barcelona, and registered at Clinicaltrials.gov (identifier: NCT02485730). All participating subjects signed the study’s informed consent form which had also been approved by the independent ethics committee ‘Parc de Salut Mar,’ Barcelona.

### 2.2. Leukocyte telomere length measurements

LTL (T/S ratio) was measured by high throughput quantitative real-time polymerase chain reaction-based assay (RT-qPCR) [35] conducted at Harvard Cancer Center Genotyping & Genetics for Population Sciences Facility. All samples were processed in a single batch, with laboratory personnel blinded to participant’s characteristics. The assay was performed in triplicate under identical conditions, with a coefficient of variation (CV) ranging between 0.15-14.6%. Samples with CV above 15% (3 samples) and those with high cycle threshold values (48 samples) were excluded from the analysis. Forty-five samples (2.6%) failed the assay due to low DNA concentration, and *APOE*-e2e4 individuals (N = 30) were also removed from the analyses. Further details can be found in [36].

### 2.3. Fluid biomarkers assessment

CSF biomarkers Aβ42, Aβ40, neurofilament light (NfL), soluble triggering receptor expressed on myeloid cells 2 (sTREM2), chitinase-3-like protein 1 (YKL40), glial fibrillary acidic protein (GFAP), S100B, neurogranin, α-synuclein, and interleukin 6 (IL-6), were measured using NeuroToolKit, a panel of exploratory robust prototype assays (Roche Diagnostics International Ltd, Rotkreuz, Switzerland) on either the Cobas® e 411 or the Cobas e 601 analyzer (Roche Diagnostics International Ltd). CSF phosphorylated tau181 (p-tau181) and total tau (t-tau) were quantified using the electrochemiluminescence Elecsys® Phospho-Tau (181P) CSF and Total-Tau CSF immunoassays (Roche Diagnostics International Ltd), respectively, on the fully automated Cobas e 601 analyzer (all Roche Diagnostics International Ltd., Rotkreuz, Switzerland), as previously described in [34]. All fluid biomarkers were measured at the Clinical Neurochemistry Laboratory, Sahlgrenska University Hospital, Mölndal, Sweden. Amyloid groups were defined with the CSF Aβ42/40 ratio (Aβ+: < 0.071). Participants were tau positive (T+) if CSF p-tau181 > 24 pg/mL or tau negative (T−) if CSF p-tau181 ≤ 24 pg/mL. Fluid biomarkers were measured at two time points: at baseline (V1) and at follow-up visits (V2). CSF biomarkers values were log-transformed to base 10. Mean (SD) follow-up time was 3.45 (0.58) years.

### 2.3. Aging and AD cortical thickness signatures

The acquisition of neuroimaging data was performed for a subset of the participants through MRI. MRI scans were obtained with a 3-tesla scanner (Ingenia CX, Philips, Amsterdam, Netherlands). The MRI protocol was identical for all participants and included a high-resolution 3D T1-weighted turbo field echo (TFE) sequence (voxel size 0.75 × 0.75 × 0.75 mm, TR/TE: 9.90/4.6 ms, flip angle = 8°). Structural T1-weighted images were segmented using FreeSurfer version 6.0 [37]. The average of the cortical thickness between hemispheres of specific brain regions was used to calculate the AD and aging brain signatures. AD brain signature was calculated as the average cortical thickness of AD-vulnerable brain regions: entorhinal, inferior temporal, middle temporal, and fusiform [38]. Aging brain signature was calculated as the average cortical thickness of aging-vulnerable brain regions: calcarine, caudal insula, cuneus, caudal fusiform, dorsomedial frontal, lateral occipital, precentral, and inferior frontal [39]. We used AD and aging brain signatures as the main outcomes to assess the association between LTL and brain structure. Higher values in these signatures represent a thicker cortex in the areas included in the signature.

### 2.4. Statistical analysis

The cross-sectional associations between LTL, CSF biomarkers and cortical thickness were assessed in the whole sample by using linear regression models. All models were adjusted by age, sex, *APOE*-4 status, as well as firmware MRI version for neuroimaging outcomes. Further, models exploring the association between LTL and the 3-year rate of change in CSF biomarker were performed. These models were additionally adjusted by the time differences between lumbar punctures. All regression β coefficients were standardized. Interactions and stratified models by *APOE*-4 and AT status were run to test differential effects by genetic AD risk and pathology. Outliers were excluded in each final sample for CSF biomarkers and LTL values, based on a threshold of 1.5 times the interquartile range. A multiple-comparison correction was applied following the Benjamini-Hochberg procedure at 5% to control for the false discovery rate (FDR). FDR was independently applied for baseline and 3-year change models, and by pre-defined CSF biomarkers pathways: amyloid pathology (i.e., Aβ42/40), neurofibrillary tangles pathology (p-tau181), neurodegeneration (i.e., NfL), synaptic dysfunction (α-synuclein, neurogranin), astrocytic response (GFAP, S100B, YKL40), microglial reactivity (sTREM2) and inflammation (IL6). A FDR-adjusted *P* value < 0.05 was considered statistically significant; unadjusted *P* value < 0.05 was considered nominally significant and unadjusted *P* value = 0.05 was considered borderline significant.

Causal mediation analysis, employing quasi-Bayesian confidence intervals with 1000 simulations was performed to assess the mediating role of CSF biomarkers in the association between LTL and neuroimaging outcomes [40]. All analyses were conducted using R software (version 4.3.3) [41].

Three datasets were used to explore the association between LTL, CSF biomarkers and neuroimaging outcomes: (1) CSF V1 dataset, including 346 individuals with available LTL and CSF biomarkers measurements obtained at V1; (2) MRI V1 dataset, including 325 individuals with available LTL, CSF biomarkers and neuroimaging data at V1; and (3) CSF longitudinal, including 237 individuals with LTL and two CSF measurements. No MRI was available for A-T+ individuals (Figure 1).

**Figure 1.**
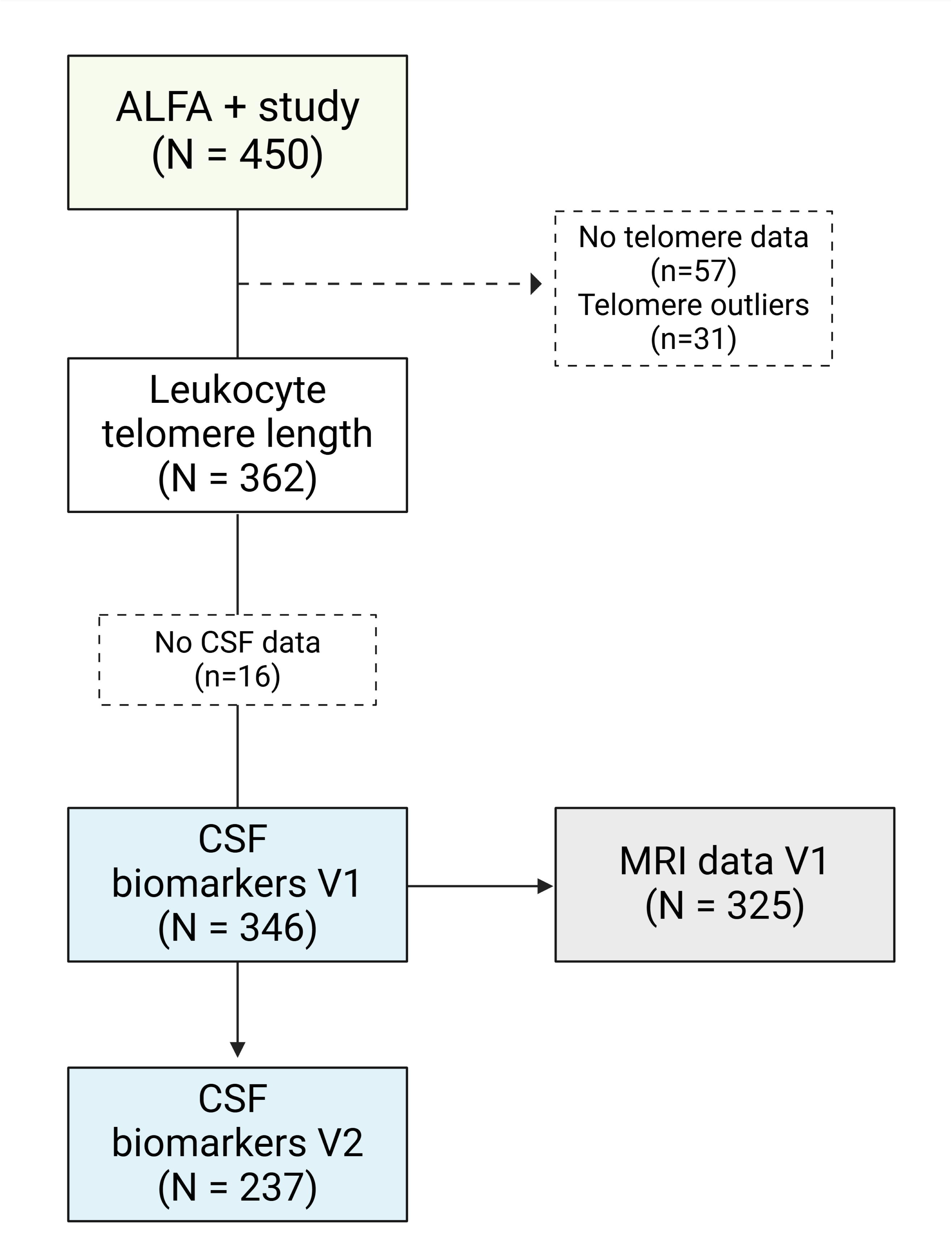
Flowchart depicting the final sample size included in the study.

## 3. Results

### 3.1. Characteristics of the study participants

Overall, no significant differences in LTL, sex, or body mass index were observed across AT stages (Table 1). However, individuals in the A+T+ group were older and had lower levels of education compared to those in the A-T-or A+T-groups. A higher proportion of *APOE*-e4 carriers was identified in the A+T-group compared to A-T- or A+T+, while no significant differences in *APOE*-e4 status were observed between the A-T- and A+T+ groups.

**Table 1.** Descriptives of study participants stratified by AT status. *Legend: 1. Median (Q1, Q3); n (%); 2. Wilcoxon rank sum test; Fisher’s exact test. Mean (SD) follow-up time was 3.45 (0.58) years*.

| Characteristics | A-T-<sup>1</sup> | N | A+T-<sup>1</sup> | N | A+T+<sup>1</sup> | N | A-T- vs.<br>A+T-<sup>2</sup> | A-T- vs.<br>A+T+<sup>2</sup> | A+T-<br>vs.<br>A+T+<sup>2</sup> | A-T+<sup>1</sup> | N |
| --- | --- | --- | --- | --- | --- | --- | --- | --- | --- | --- | --- |
| Leukocyte telomere length (T/S) | 1.03 (0.92, 1.17) | 215 | 1.02 (0.90, 1.15) | 92 | 1.04 (0.99, 1.12) | 26 | 0.687 | 0.841 | 0.685 | 1.02 (0.92, 1.06) | 13 |
| Age (years) | 60.2 (57.0, 64.0) | 215 | 61.9 (58.2, 65.2) | 92 | 64.1 (62.5, 66.6) | 26 | 0.147 | <0.001 | 0.025 | 62.6 (60.4, 63.2) | 13 |
| Females, n (%) | 136 (63%) | 215 | 52 (57%) | 92 | 18 (69%) | 26 | 0.307 | 0.667 | 0.268 | 9 (69%) | 13 |
| Education (years) | 12 (11, 17) | 215 | 14 (11, 17) | 92 | 11 (8, 17) | 26 | 0.930 | 0.025 | 0.036 | 17 (12, 17) | 13 |
| Body mass index (kg/m<sup>2</sup>) | 26.4 (24.5, 30.1) |  | 26.2 (23.9, 28.8) |  | 26.3 (24.3, 28.6) |  | 0.468 | 0.306 | 0.583 | 26.2 (22.9, 28.0) |  |
| **APOE-e4 carriers, n (%)** | 88 (41%) | 215 | 76 (83%) | 92 | 15 (58%) | 26 | <0.001 | 0.141 | 0.015 | 5 (38%) | 13 |
| **CSF biomarkers at V1** |  |  |  |  |  |  |  |  |  |  |  |
| Aβ42, (pg/mL) | 1,397 (1,085, 1,819) | 215 | 801 (669, 992) | 92 | 842 (720, 1,189) | 26 | <0.001 | <0.001 | 0.123 | 2,575 (2,113, 2,803) | 13 |
| Aβ40, (pg/mL) | 16,530 (13,380, 20,060) | 215 | 15,220 (13,490, 17,370) | 92 | 22,505 (19,080, 25,030) | 26 | 0.028 | <0.001 | <0.001 | 26,010 (25,980, 27,920) | 13 |
| Aβ42/40 ratio | 0.085 (0.081, 0.093) | 215 | 0.054 (0.044, 0.064) | 92 | 0.042 (0.031, 0.051) | 26 | <0.001 | <0.001 | <0.001 | 0.098 (0.091, 0.104) | 13 |
| p-tau181, (pg/mL) | 13 (10, 17) | 215 | 15 (12, 18) | 92 | 29 (27, 33) | 26 | 0.007 | <0.001 | <0.001 | 25 (24, 29) | 13 |
| NfL, (pg/mL) | 74 (59, 88) | 215 | 80 (63, 95) | 92 | 111 (93, 138) | 26 | 0.033 | <0.001 | <0.001 | 95 (85, 100) | 13 |
| t-tau, (pg/mL) | 169 (139, 209) | 215 | 183 (153, 218) | 92 | 323 (302, 364) | 26 | 0.029 | <0.001 | <0.001 | 303 (287, 325) | 13 |
| neurogranin, (pg/mL) | 680 (540, 895) | 215 | 686 (571, 868) | 92 | 1,314 (1,178, 1,443) | 26 | 0.605 | <0.001 | <0.001 | 1,356 (1,283, 1,419) | 13 |
| α-synuclein, (pg/mL) | 173 (138, 230) | 215 | 170 (146, 207) | 92 | 295 (244, 352) | 26 | 0.605 | <0.001 | <0.001 | 311 (298, 356) | 13 |
| GFAP, (pg/mL) | 6,940 (5,700, 8,700) | 215 | 7,685 (5,800, 8,720) | 92 | 10,755 (9,050, 11,910) | 26 | 0.524 | <0.001 | <0.001 | 7,510 (6,310, 9,900) | 13 |
| S100B, (pg/mL) | 966 (852, 1,120) | 215 | 1,030 (832, 1,210) | 92 | 1,130 (892, 1,260) | 26 | 0.117 | 0.003 | 0.073 | 1,120 (869, 1,210) | 13 |
| YKL-40, (pg/mL) | 133,300 (105,400, 159,300) | 215 | 137,400 (100,950, 166,750) | 92 | 200,850 (174,800, 234,100) | 26 | 0.745 | <0.001 | <0.001 | 201,800 (150,000, 217,600) | 13 |
| sTREM2, (pg/mL) | 7,550 (6,380, 8,950) | 215 | 7,185 (5,960, 8,740) | 92 | 9,875 (8,700, 11,480) | 26 | 0.141 | <0.001 | <0.001 | 10,250 (9,930, 13,480) | 13 |
| IL-6, (pg/mL) | 3.57 (2.91, 4.41) | 215 | 3.54 (2.85, 4.44) | 92 | 3.40 (2.77, 4.63) | 26 | 0.912 | 0.891 | 0.755 | 2.98 (2.42, 4.65) | 13 |
| **CSF biomarkers at V2** |  |  |  |  |  |  |  |  |  |  |  |
| Aβ42, (pg/mL) | 1,424 (1,104, 1,893) | 147 | 755 (676, 989) | 61 | 828 (661, 1,128) | 17 | <0.001 | <0.001 | 0.717 | 2,249 (1,855, 3,382) | 12 |
| Aβ40, (pg/mL) | 16,740 (13,690, 20,860) | 147 | 16,250 (14,360, 19,350) | 61 | 22,410 (18,830, 25,780) | 17 | 0.725 | <0.001 | <0.001 | 25,675 (23,565, 28,975) | 12 |
| Aβ42/40 ratio | 0.088 (0.077, 0.098) | 147 | 0.048 (0.040, 0.060) | 61 | 0.038 (0.032, 0.049) | 17 | <0.001 | <0.001 | 0.003 | 0.098 (0.073, 0.121) | 12 |
| p-tau181, (pg/mL) | 15 (12, 19) | 147 | 17 (15, 21) | 61 | 32 (29, 39) | 17 | <0.001 | <0.001 | <0.001 | 26 (22, 29) | 12 |
| NfL, (pg/mL) | 93 (73, 117) | 147 | 101 (78, 123) | 61 | 138 (119, 180) | 17 | 0.152 | <0.001 | <0.001 | 104 (97, 126) | 12 |
| t-tau, (pg/mL) | 175 (145, 225) | 147 | 201 (169, 260) | 61 | 336 (292, 443) | 17 | <0.001 | <0.001 | <0.001 | 300 (260, 337) | 12 |
| neurogranin, (pg/mL) | 730 (566, 1,018) | 147 | 837 (724, 1,092) | 61 | 1,443 (1,347, 1,623) | 17 | 0.016 | <0.001 | <0.001 | 1,412 (1,199, 1,521) | 12 |
| α-synuclein, (pg/mL) | 181 (131, 243) | 147 | 212 (158, 251) | 61 | 322 (275, 403) | 17 | 0.079 | <0.001 | <0.001 | 312 (281, 360) | 12 |
| GFAP, (pg/mL) | 9,790 (7,840, 12,040) | 147 | 10,320 (8,250, 12,550) |  | 15,080 (14,070, 16,870) |  | 0.665 | <0.001 | <0.001 | 11,320 (8,805, 12,855) | 12 |
| S100B, (pg/mL) | 964 (834, 1,090) | 147 | 1,010 (867, 1,150) | 61 | 1,160 (898, 1,320) | 17 | 0.271 | 0.007 | 0.098 | 1,155 (1,000, 1,195) | 12 |
| YKL-40, (pg/mL) | 149,300 (117,800, 186,100) | 147 | 163,100 (135,300, 195,900) | 61 | 229,000 (198,200, 272,500) | 17 | 0.098 | <0.001 | <0.001 | 218,300 (167,750, 283,500) | 12 |
| sTREM2, (pg/mL) | 8,900 (7,480, 10,530) | 147 | 9,280 (7,390, 10,420) | 61 | 10,830 (9,200, 14,000) | 17 | 0.834 | 0.006 | 0.010 | 12,945 (11,950, 16,090) | 12 |
| IL-6, (pg/mL) | 4.26 (3.58, 5.29) | 147 | 4.40 (3.56, 5.25) | 61 | 4.03 (3.38, 5.07) | 17 | 0.703 | 0.571 | 0.417 | 3.76 (3.12, 3.89) | 12 |
| **Cortical thickness** |  |  |  |  |  |  |  |  |  |  |  |
| Aging signature | 2.28 (2.23, 2.35) | 209 | 2.28 (2.25, 2.34) | 91 | 2.26 (2.18, 2.31) | 25 | 0.362 | 0.075 | 0.036 | - | 0 |
| AD signature | 2.53 (2.46, 2.58) | 209 | 2.54 (2.48, 2.60) | 91 | 2.47 (2.42, 2.57) | 25 | 0.129 | 0.055 | 0.008 | - | 0 |

1 Median (Q1, Q3); n (%)
Mean (SD) follow-up time was 3.45 (0.58) years.

CSF concentrations of NfL and t-tau were elevated across AT stages at V1. At V2, no significant differences in CSF NfL concentrations were found between A-T- and A+T-. CSF concentrations of neurogranin, α-synuclein, GFAP, S100B, YKL-40, and sTREM2 were significantly higher in A+T+ compared to A-T-, with similar elevations (excluding S100B) noted in A+T+ compared to A+T-. In contrast, no significant differences in these biomarker concentrations were observed between A-T- and A+T-at either visit. CSF IL-6 concentrations did not differ across AT stages at V1 or V2. Moreover, at V1, individuals in the A+T+ group exhibited thinner brain cortex compared to the A+T-group for both aging- and AD-vulnerable brain regions, with a trend toward thinner cortices also observed in A+T-individuals compared to A-T-.

When stratified by *APOE*-e4 status, we observed higher proportion of AT-positive individuals and lower CSF sTREM2 concentrations among e4 carriers, at both at V1 and V2. Higher cortical thickness was observed among *APOE*-e4 carriers in AD vulnerable regions (Table S1 in supporting information).

### 3.2. Association between LTL and AD-related CSF biomarkers

In the whole sample shorter LTL was associated with higher baseline GFAP (β = −0.11, P = 0.042) (Figure 2A). This association remained statistically significant after adjusting for CSF Aβ42/40 (β = −0.11, P = 0.044). However, the association was no longer statistically significant after controlling for CSF p-tau181 (β = −0.08, P = 0.103), or Aβ40 levels which account for differences in CSF production and clearance rates (β = −0.08, P = 0.084) [42]. Similarly, shorter LTL was associated with higher baseline levels of S100B (β = −0.13, P = 0.013) (Figure 2B). This association persisted after adjusting for CSF Aβ42/40 ratio (β = −0.13, P = 0.014), p-tau181 (β = −0.12, P = 0.027), and Aβ40 levels (β = −0.12, P = 0.024).

**Figure 2.**
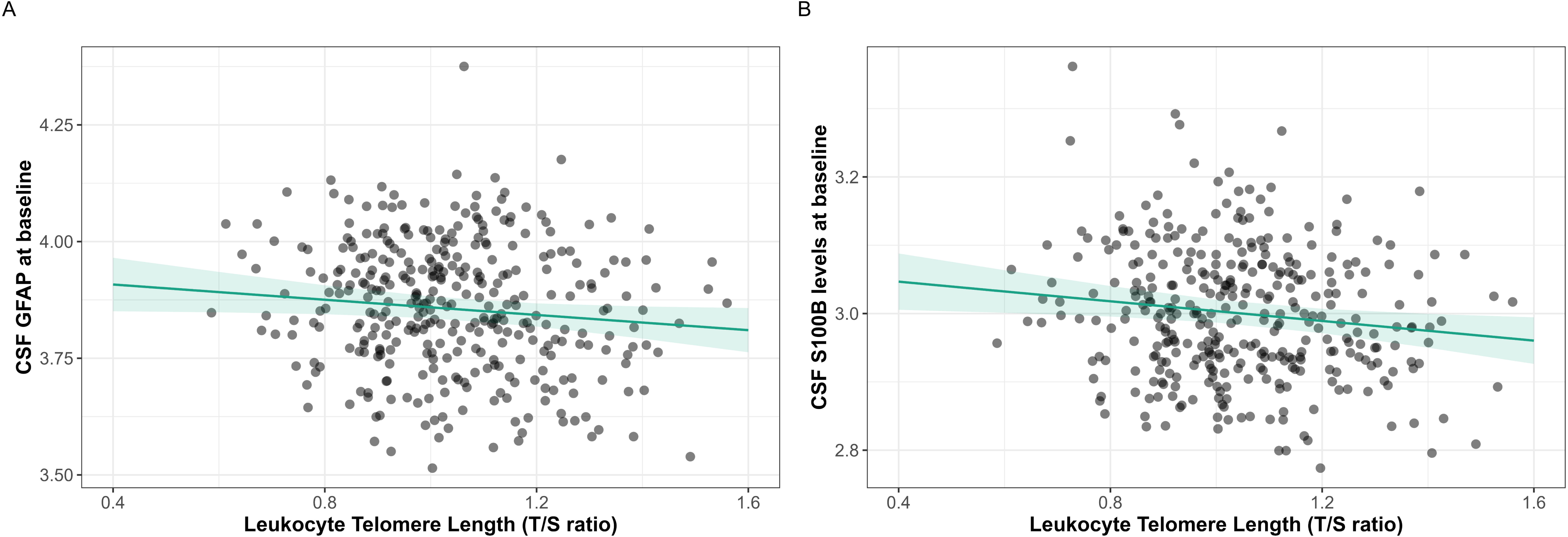
Significant associations between LTL and baseline log-transformed (A) CSF GFAP and (B) CSF S100B levels in the global sample (N = 346). *Models were adjusted for age, sex, and APOE-e4 status*.

Longitudinally, shorter LTL was associated with increased CSF α-synuclein levels over time (β = −0.14, P = 0.042) (Figure 3). This association remained statistically significant after adjusting for CSF Aβ42/40 ratio (β = −0.14, P = 0.042), p-tau181 (β = −0.13, P = 0.044), and Aβ40 baseline levels (β = −0.13, P = 0.046).

**Figure 3.**
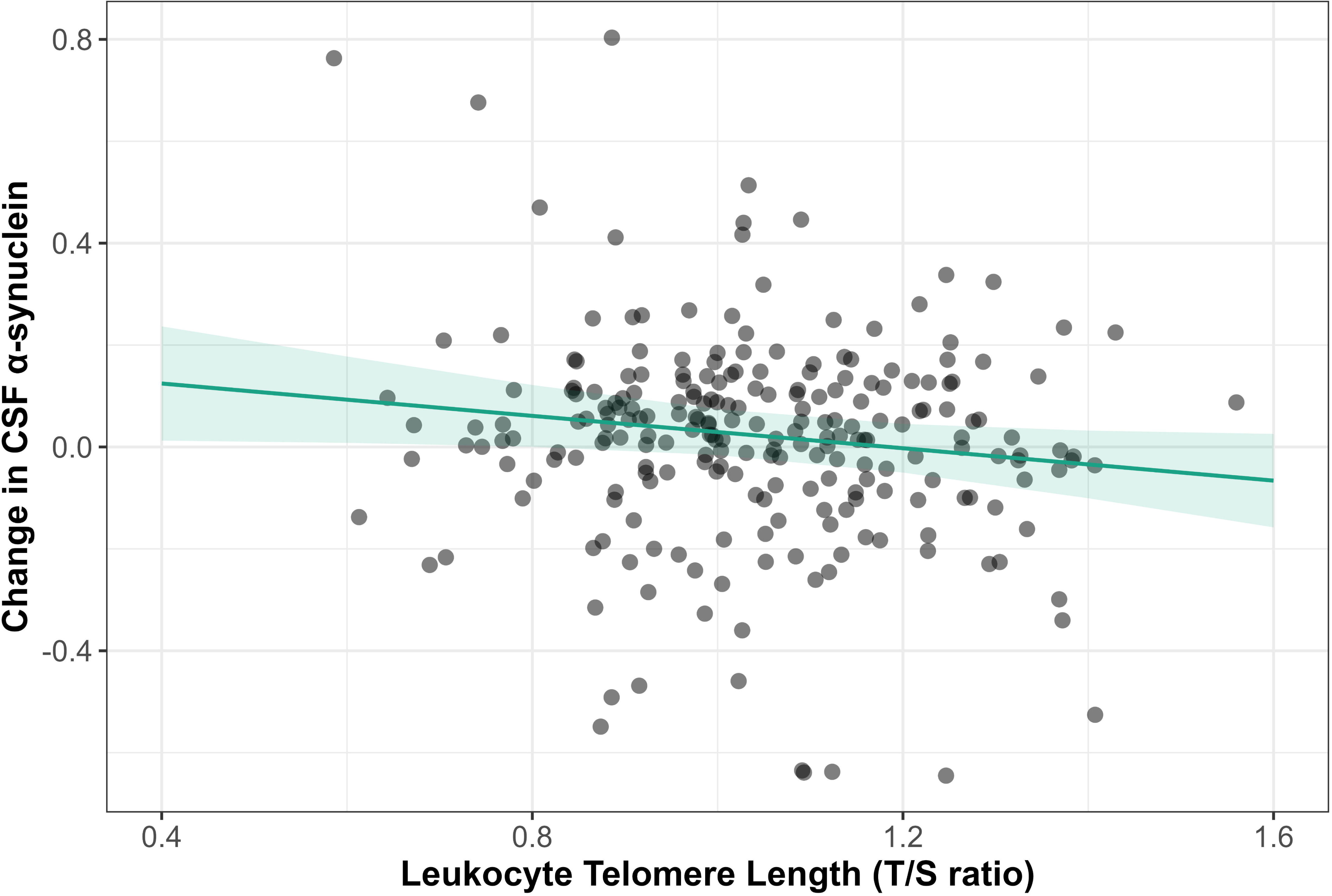
Significant longitudinal association between LTL and log-transformed change in CSF α-synuclein levels over time (Two-time-points sample N = 237). *Model was adjusted for age, sex, APOE-e4 status, and time difference between visits*.

No other statistically significant associations were found in the whole sample (Table S2).

### 3.2. Modification by APOE-e4 status on CSF biomarkers

LTL showed statistically significant interactions with *APOE*-e4 status on the association with CSF YKL-40 at baseline (β*int* = −0.2, P*int* = 0.047) and the change in CSF sTREM2 over time (β*int* = 0.27, P*int* = 0.036). Specifically, shorter LTL was associated with higher baseline CSF YKL-40 (β = −0.19, P = 0.006) and decreasing CSF sTREM2 over time (β = 0.17, P = 0.046) only among e4 carriers (Figures S1-S2). No other statistically significant interactions between LTL and *APOE*-e4 status were observed (Table S3 in supporting information).

Nonetheless, when stratifying by *APOE*-e4 status (Figure 4), shorter LTL was associated with higher p-tau181 (β = −0.15, P = 0.033), NfL (β = −0.15, P = 0.026), t-tau (β = −0.17, P = 0.021), neurogranin (β = −0.15, P = 0.034), GFAP (β = −0.15, P = 0.040) and YKL-40 at baseline in carriers of the e4 allele. In addition, shorter LTL was associated with longitudinal increases of α-synuclein (β = −0.18, P = 0.05) and decreases of sTREM2 over time (β = 0.17, P = 0.046) among *APOE*-e4 carriers.

**Figure 4.**
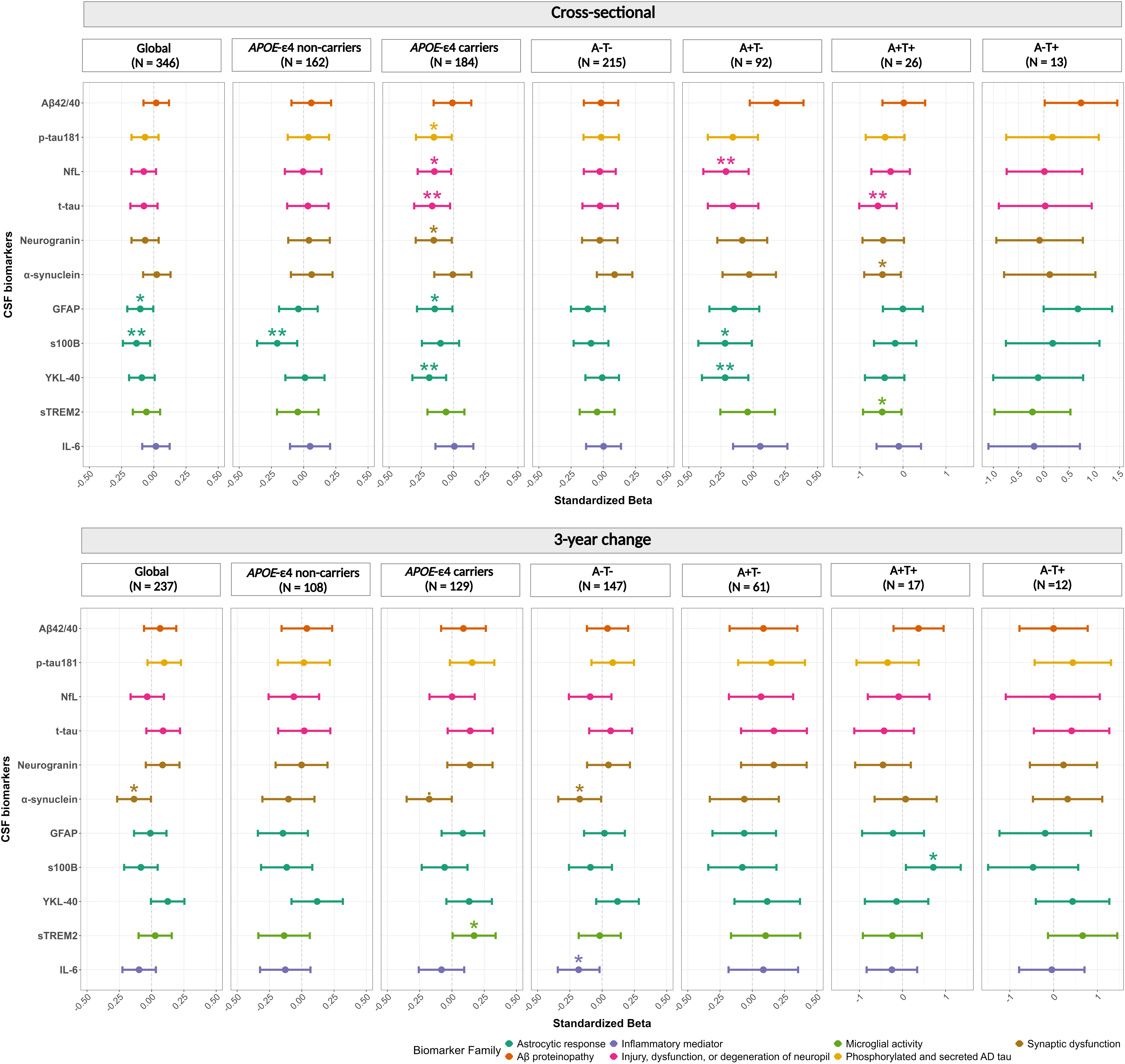
Significant association between leukocyte telomere length and log-transformed CSF biomarkers in the global sample and stratified by *APOE*-e4 and AT status. *Each point represents the standardized beta for one association model, horizontal lines represent confidence intervals and colors represent the biomarker family for multiple comparison correction). Cross-sectional models were adjusted for age, sex, and APOE-e4 status, while longitudinal models were further adjusted for time between visits*.

In contrast, among *APOE*-e4 non-carriers, shorter LTL was associated with higher baseline S100B (β = −0.21, P = 0.010) (Table S4, Figure S3 in supporting information). These associations were independent of Aβ status, while trends were observed for CSF NfL, t-tau and GFAP, only the association with YKL-40 remained statistically significant after adjusting for T status (Tables S5-S6 in supporting information).

### 3.3. Modification by AT status on CSF biomarkers

LTL showed significant interactions with AT status in its association with baseline CSF Aβ42/40 (A-T+ vs. A-T-: β*int* = 3.89, P*int* = 0.002; A+T-vs. A-T-: β*int* = 0.57, P = 0.09) (Figure S4 in supporting information). When stratifying by AT status (Figure 4), trends were observed between LTL and CSF Aβ42/40 in both A-T+ individuals (β = 0.74, P = 0.080) and A+T-individuals (β = 0.18; P = 0.091). Regarding p-tau pathology, no significant interactions or associations were found in any AT groups. However, a trend was observed among A+T+ individuals with a suggestive negative association between LTL and CSF p-tau181 (β = −0.42; P = 0.085). Even though no other interactions between LTL and AT status were observed statistically significant associations were found between LTL and CSF biomarkers when stratifying by AT status (Tables S4, S7 in supporting information).

In A-T-individuals, shorter LTL was longitudinally associated with increased CSF α-synuclein (β = −0.17, P = 0.042) and IL-6 over time (β = −0.18, P = 0.029) (Figure S5 in supporting information). In the A+T-group, shorter LTL was associated with higher baseline CSF NfL (β = −0.21, P = 0.021), S100B (β = - 0.22, P = 0.042) and YKL-40 (β = −0.22, P = 0.020), whereas no significant associations were found for the change in CSF concentrations over time (Figure S6). Among A+T+ individuals, shorter LTL also associated with higher CSF t-tau (β = −0.58, P = 0.016), α-synuclein (β = −0.48, P = 0.039), sTREM2 (β = - 0.49, P = 0.046) at baseline. In addition, shorter LTL was longitudinally associated with decreased CSF S100B levels over time among A+T+ individuals (β = 0.72, P = 0.049) (Figure S7). No significant associations were observed in the A-T+ group.

### 3.4. Imaging biomarkers and leukocyte telomere length

Shorter LTL was associated with a thicker brain cortex in regions vulnerable to AD-related neurodegeneration (β = −0.11, P = 0.046) (Figure 5A). This association was independent of CSF Aβ42/40 (β = −0.11, P = 0.046), p-tau181 (β = −0.12, P = 0.035), and NfL (β = −0.13, P = 0.021). Shorter LTL was associated with thicker cortex in aging-vulnerable brain regions (β = −0.13, P = 0.019) (Figure 5B). This association was independent of CSF Aβ42/40 (β = - 0.13, P = 0.019), p-tau181 (β = −0.13, P = 0.019), and NfL (β = −0.13, P = 0.017) (Table S8).

**Figure 5.**
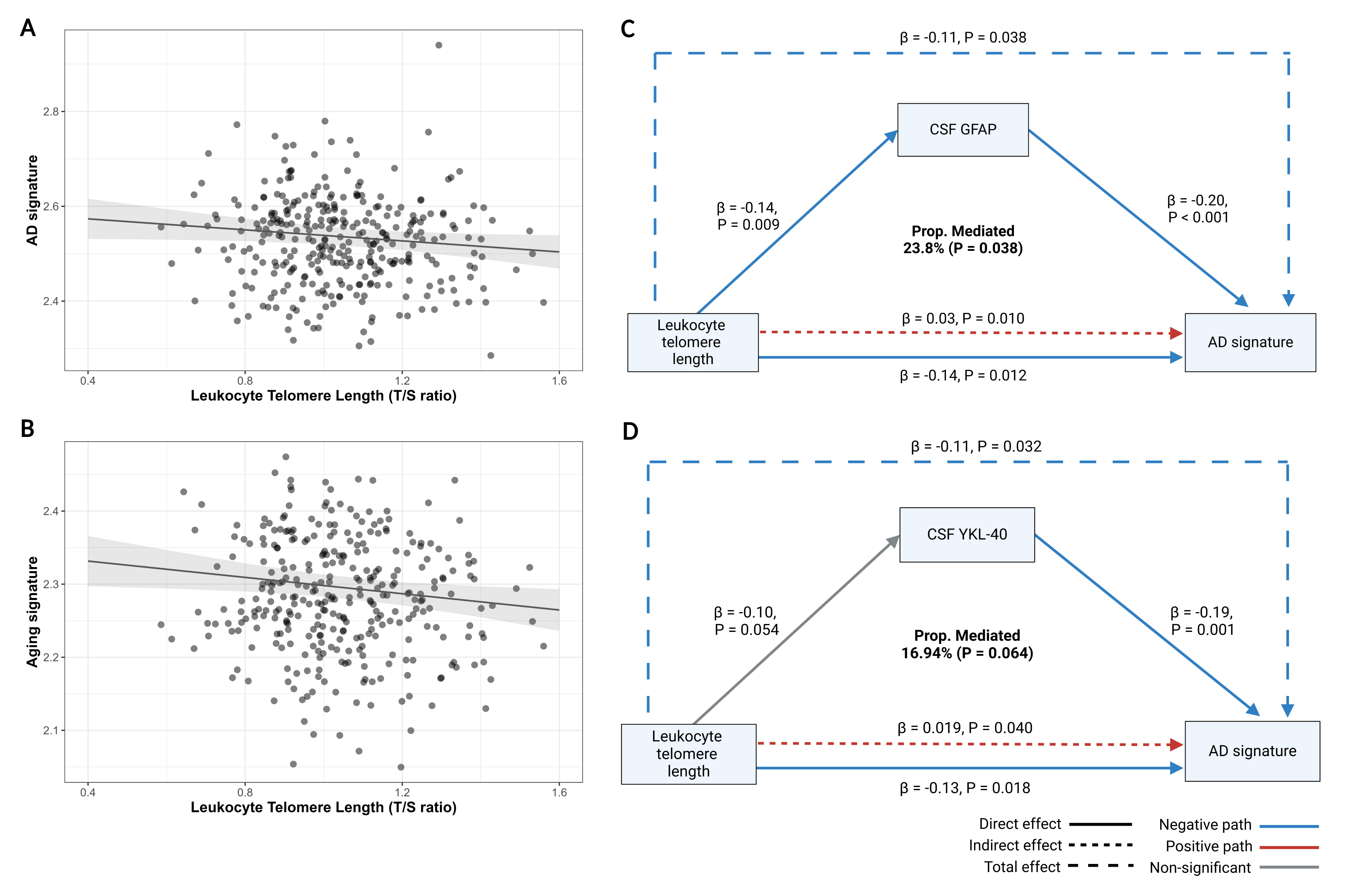
Significant associations between leukocyte telomere length and cortical thickness in the (A) AD signature (B) aging signature. Mediator role of (C) CSF GFAP and (D) CSF YKL-40 on the association between leukocyte telomere length and AD cortical thickness signatures (N = 325). *Models were corrected for age, sex, APOE-e4 status, and firmware MRI version. All coefficients are standardized*.

Previous studies have reported positive associations between CSF biomarkers of glial reactivity and inflammation with higher gray matter volumes and thicker cortical thickness [43–45]. Therefore, we investigated whether shorter LTL’s association with increasing CSF glial and inflammatory biomarkers could mediate its association with cortical thickness. Specifically, we explored the putative mediating role of glial (i.e., GFAP, S100B, YKL-40 and sTREM2) and inflammatory biomarkers (i.e. IL-6) in the association between LTL and cortical thickness.

The association between shorter LTL and higher cortical thickness in AD-vulnerable regions was partially mediated by GFAP (i.e., 23.8% of the effect mediated) (P = 0.038). Specifically, shorter LTL was indirectly associated with thinner cortex in AD signature through its effect on GFAP (Indirect pathway: β = 0.03, P = 0.010). However, the direct negative association between LTL and AD signature was still significant after accounting for the GFAP pathway (Direct pathway: β = −0.14, P = 0.012) (Figure 5C).

In addition, a significant indirect association between shorter LTL and thinner AD signature through YKL-40 was observed (Indirect pathway: β = 0.019, P = 0.040). The direct negative association persisted after accounting for the YKL-40 pathway (Direct pathway: β = −0.13, P = 0.018). A borderline mediating role of YKL-40 in the association between LTL and AD signature was observed (Mediation proportion: 16.94%; P = 0.064) (Figure 5D).

No evidence for other mediation was detected for CSF S100B, sTREM2 or IL-6 in the association between LTL and AD signature. Regarding the aging signature, a borderline significant mediating effect was observed for YKL-40 (Mediation proportion 16.19%, P = 0.088). LTL was indirectly and positively associated with the aging signature through YKL-40 (Indirect pathway: β = 0.02, P = 0.036). Nonetheless, shorter LTL remained associated with higher cortical thickness in aging-vulnerable regions after accounting for YKL-40 indirect pathway (Direct pathway: β = −0.13, P = 0.018). No evidence for other mediating roles were detected for CSF GFAP, S100B, sTREM2 or IL-6 in the association between LTL and aging signature (Figure S8 in supporting information).

## 4. Discussion

In this study, we investigated the association between LTL, AD-related CSF biomarkers (baseline and 3-year changes), and cortical thickness in CU individuals at increased risk of AD. We showed that shorter LTL was associated with higher levels of astrocytic reactivity and synaptic dysfunction biomarkers, as well as thicker cortex in aging and AD-vulnerable brain regions. Notably, astrocytic biomarkers partially mediated the relationship between LTL and cortical thickness, suggesting a potential pathway through which telomere shortening may influence early neurodegeneration in AD. Additionally, the relationship between LTL and AD-related CSF biomarkers differed according to *APOE*-e4 and AT status.

To our knowledge, this is the first study to comprehensively examine the relationship between LTL and an extensive set of AD-related CSF biomarkers. In the whole sample, no significant associations were found between LTL and CSF Aβ or p-tau biomarkers, consistent with previous studies focused on cognitively impaired patients with MCI or AD [23,46]. Unlike prior work, our study accounted for the modifying effects of *APOE*-e4 and AD pathology in preclinical stages, revealing that shorter LTL was significantly associated with higher baseline p-tau181 levels in *APOE*-e4 carriers. Although no significant associations were observed when stratifying by AT status for p-tau181, a trend of increasingly negative coefficients across AT stages suggests that the association with p-tau181 in e4 carriers may be influenced by the progression of the disease, potentially diluted by smaller sample sizes in later stages. Similarly, shorter LTL was associated to increased neurodegeneration biomarkers in *APOE*-e4 carriers, including NfL and t-tau, primarily in A+T- and A+T+ individuals, respectively.

Shorter LTL was associated with higher baseline CSF α-synuclein in A+T+ individuals and an increase in α-synuclein over time in the overall sample and A-T-individuals. As we measured total CSF α-synuclein, this likely reflects the effect of shorter LTL on synaptic loss and neuronal injury [34]. Previous studies show that shorter telomeres induce aging-related phenotypes in iPSC-derived neurons, such as reduced neurite density and length [47], though the role of telomeres in neuronal maintenance and synaptic plasticity in humans is not well understood. Additionally, shorter LTL was associated with higher CSF neurogranin concentrations in *APOE*-e4 carriers. Neurogranin, a postsynaptic protein important for memory formation [48,49], is associated with cognitive resilience during aging and increases in the CSF of AD patients due to synaptic degeneration [50–56]. Genes involved in telomere maintenance have pleiotropic effects on brain expression, methylation, and chromatin profiles with effects on synapsis homeostasis [57]. Given that LTL is highly heritable and partially determined early in life [58,59], the association between LTL and synaptic biomarkers may reflect the pleiotropic roles of telomere maintenance pathways in synaptic plasticity during brain development and aging. These effects may be amplified under pathological conditions, particularly in *APOE*-e4 carriers, due to its role in synapse pruning and neuroinflammation [60].

Several studies have reported increased CSF glial biomarkers across the AD *continuum* [34,61–66]. In our study, LTL significantly interacted with *APOE*-e4 status in the association with CSF YKL-40, with shorter LTL linked to higher baseline YKL-40 levels exclusively in *APOE*-e4 carriers. YKL-40 is a glycoprotein secreted by multiple cell-types, including leukocytes and endothelial cells, which participates in systemic inflammation and angiogenesis [67,68]. In the brain, YKL-40 is highly expressed in astrocytes associated with Aβ deposits [69] and increased CSF YKL-40 concentrations are found since prodromal AD [70]. In our sample, CSF YKL-40 was previously linked to tau phosphorylation and Aβ-related neuronal injury [71]. In addition, longitudinally, a significant interaction between LTL and *APOE*-e4 status was observed with the change in CSF sTREM2 over time. Shorter LTL was longitudinally associated with decreasing CSF sTREM2 among *APOE*-e4 carriers. This is in line with the notion of an impaired microglial-response to injury in aging and among *APOE*-e4 carriers [72,73]. However, shorter LTL was associated with higher CSF sTREM2 at baseline in A+T+ individuals. Elevated CSF sTREM2 have been observed in the early stages of AD after the onset of amyloid pathology [61,74–77]. In individuals at the preclinical stage of sporadic AD, higher baseline CSF sTREM2 was linked to a deceleration in tau deposition, as measured by tau-PET [78]. On the contrary, increases in CSF sTREM2 mediated amyloid-related p-tau181 pathology at the earliest stages of the AD *continuum* [79]. In our sample, higher baseline CSF sTREM2 was associated with better memory and executive outcomes, independent of AD pathology [80]. The contrasting baseline and longitudinal associations between LTL and CSF sTREM2 suggest a complex interplay between LTL, microglial activation, and potentially neuroinflammation across the disease *continuum*. These findings align with previous studies linking longer LTL to worse cognitive outcomes in biomarker-positive individuals [23]. Further longitudinal studies, including individuals at more advanced biological and clinical stages of AD, are needed to clarify LTL’s role in microglial reactivity.

Additionally, shorter LTL was associated with higher CSF GFAP levels in the whole sample, an association primarily driven by *APOE*-e4 carriers. A similar trend was observed in the A-T-group but diminished across AT stages, potentially due to reduced statistical power with smaller sample sizes. The high proportion of *APOE*-e4 carriers in the A+T-group (i.e., 81%) suggests LTL may influence astrocytic reactivity through an ApoE-related pathway early in the AD *continuum*, even before reaching the CSF Aβ positivity threshold. Additionally, shorter LTL was associated with higher baseline CSF S100B in *APOE*-e4 non-carriers and A+T-individuals, but with longitudinal decreases in CSF S100B in A+T+ individuals. S100B is a calcium-binding protein primarily expressed in astrocytes in the brain and acts as a damage-associated molecular pattern, released under cell stress [81]. S100B is found to be elevated in CSF across various neurodegenerative diseases, including AD, and correlates with brain atrophy and cognitive decline [82–84]. Finally, shorter LTL was associated with increased CSF IL-6 over time in A-T-individuals.

Together, these findings suggest that telomere maintenance may influence glial activity and neuroinflammation in an ApoE and pathology-associated manner, particularly in early AD stages. Interestingly, shorter telomeres in human iPSC-derived astrocytes from old donors were related to increased production of inflammatory cytokines, GFAP expression and elevated DNA damage when compared to younger donors [47]. In addition, distinct inflammatory and glial signatures have been found in relation to *APOE* genotype and cognitive performance stages in AD patients [85], suggesting that LTL could play a role in pathways associated with resilience to disease progression.

We observed an unexpected negative association between LTL and cortical thickness in regions vulnerable to aging and AD. This contrasts with prior studies linking longer LTL to greater gray matter volumes and cortical thickness [19,86,87]. However, previous studies have described transient increases in cortical thickness in AD-susceptible regions among CU individuals at risk for AD, primarily attributed to early gliosis and Aβ pathology accumulation [45,88–92]. In line with this, our sample previously showed associations between higher CSF glial and inflammatory biomarker concentrations and increased cortical volumes [43,44]. In the current study, the association between LTL and cortical thickness was independent of CSF Aβ42/40, p-tau181, and NfL, and partially mediated by astrocytic biomarkers, including GFAP and YKL-40. Interestingly, while shorter LTL showed an indirect positive association with cortical thickness through these biomarkers, the direct association between shorter LTL and increased cortical thickness persisted even after accounting for these mediators.

The mechanisms by which telomere shortening impacts brain structure remain unclear [87], but may involve cellular aging, oxidative stress, and immune dysregulation [93,94]. LTL is inversely and strongly correlated with a blood proteomic age clock, primarily driven by immune and inflammatory cytokines, which is linked to mortality, cognitive decline, and neurodegeneration [95]. Blood leukocyte telomeres serve as a marker of peripheral immune aging, associated with chronic systemic inflammation and increased infection risk [7,96,97]. Growing evidence suggests that heightened systemic inflammation and activation of the peripheral innate immune system can trigger microglial activation and neuroinflammation in aging and neurodegeneration [98]. Based on our findings, we might speculate that peripheral immune aging, represented by leukocyte telomere shortening, may influence glial reactivity via the neuroimmune axis, disrupting brain microstructure homeostasis as reflected in cortical thickness [99–101]. Additionally, LTL’s dynamic effects on glial activation in the context of AD pathology may underlie its non-linear relationship with cognitive performance and AD risk across the disease *continuum* [22,36,102], warranting further investigation.

Our study is not without limitations. The ALFA+ cohort includes middle-aged, CU individuals at increased risk of AD, which may limit generalizability to the general population, particularly to biomarker-positive individuals at later AD stages or with comorbidities. The 3.5-year follow-up period also restricted our ability to capture long-term biomarker trajectories. Finally, the relatively modest sample size may have constrained our power to detect associations surviving multiple comparison corrections, but the observed trends warrant further investigation in larger cohorts with multimodal biomarker data.

In conclusion, our findings highlight the role of LTL in AD pathophysiology, particularly in synaptic dysfunction and glial homeostasis. Importantly, astrocytic reactivity mediated the relationship between LTL and cortical thickness, suggesting a pathway linking peripheral biological aging to neurodegenerative processes in the brain. Further research is needed to clarify the mechanisms underlying LTL’s role in neurodegeneration, with the potential to identify determinants of resilience and resistance to pathology leading to successful cognitive aging.

## Data Availability

All data produced in the present study are available upon reasonable request to the authors.

## ACKNOWLEDGMENTS

This publication is part of the ALFA study (ALzheimers and Families). The authors would like to thank the collaborators of the ALFA study. Additionally, the authors would like to express their most sincere gratitude to the ALFA project participants and relatives without whom this research would not have been possible. The authors thank Roche Diagnostics International Ltd for providing the kits to measure CSF biomarkers. The Roche NeuroToolKit is a panel of exploratory prototype assays designed to robustly evaluate biomarkers associated with key pathologic events characteristic of AD and other neurological disorders, used for research purposes only and not approved for clinical use. COBAS and ELECSYS are trademarks of Roche. All other product names and trademarks are the property of their respective owners. The ALFA+ study receives funding from “la Caixa” Foundation (ID 100010434), under agreement LCF/PR/GN17/50300004 and the Alzheimer’s Association and an international anonymous charity foundation through the TriBEKa Imaging Platform project (TriBEKa17519007). Additional support has been received from the Universities and Research Secretariat, Ministry of Business and Knowledge of the Catalan Government under the grant no. 2021 SGR 00913. CM received funding within the context of EURO-FINGERS, a EU Joint Programme – Neurodegenerative Disease Research (JPND) project. HZ is a Wallenberg Scholar and a Distinguished Professor at the Swedish Research Council supported by grants from the Swedish Research Council (#2023-00356, #2022-01018 and #2019-02397), the European Union’s Horizon Europe research and innovation programme under grant agreement No 101053962, Swedish State Support for Clinical Research (#ALFGBG-71320), the Alzheimer Drug Discovery Foundation (ADDF), USA (#201809-2016862), the AD Strategic Fund and the Alzheimer’s Association (#ADSF-21-831376-C, #ADSF-21-831381-C, #ADSF-21-831377-C, and #ADSF-24-1284328-C), the European Partnership on Metrology, co-financed from the European Union’s Horizon Europe Research and Innovation Programme and by the Participating States (NEuroBioStand, #22HLT07), the Bluefield Project, Cure Alzheimer’s Fund, the Olav Thon Foundation, the Erling-Persson Family Foundation, Familjen Rönströms Stiftelse, Stiftelsen för Gamla Tjänarinnor, Hjärnfonden, Sweden (#FO2022-0270), the European Union’s Horizon 2020 research and innovation programme under the Marie Skłodowska-Curie grant agreement No 860197 (MIRIADE), the European Union Joint Programme – Neurodegenerative Disease Research (JPND2021-00694), the National Institute for Health and Care Research University College London Hospitals Biomedical Research Centre, and the UK Dementia Research Institute at UCL (UKDRI-1003). KB is supported by the Swedish Research Council (#201700915); the Alzheimer Drug Discovery Foundation (ADDF), USA (#RDAPB2018092016615); the Swedish Alzheimer Foundation (#AF742881); Hjärnfonden, Sweden (#FO20170243); the Swedish state under the agreement between the Swedish government and the County Councils, the ALFagreement (#ALFGBG715986); the European Union Joint Program for Neurodegenerative Disorders (JPND2019466236); the National Institute of Health (NIH), USA (grant #1R01AG06839801); and the Alzheimer’s Association 2021 Zenith Award (ZEN21848495). MSC receives funding from the European Research Council (ERC) under the European Union’s Horizon 2020 research and innovation program (Grant agreement No. 948677), the Instituto de Salud Carlos III (ISCIII) through the projects PI19/00155 and PI22/00456 (Co-funded by European Regional Development Fund (FEDER) “A way to make Europe”), and receives the support of a fellowship funded by “la Caixa” Foundation (ID 100010434), and the European Union’s Horizon 2020 research and innovation programme under the Marie Skłodowska-Curie grant agreement No 847648 (fellowship code LCF/BQ/PR21/11840004). OGR receives funding from the Alzheimer’s Association Research Fellowship Program (2019-AARF-644568), from Instituto de Salud Carlos III (ISCIII) through the project PI19/00117 co-funded by the European Union (FEDER), and from Spanish Ministry of Science and Innovation - State Research Agency MCIN/AEI/10.13039/501100011033 through the project IJC2020-043417-I, co-funded by the European Union “Next GenerationEU”/PRTR. NVT is supported by the Spanish Ministry of Science and Innovation - State Research Agency MCIN/AEI/10.13039/501100011033 through the project IJDC2020-043216-I, co-funded by the European Union “Next GenerationEU”/PRTR), and project PID2022-143106OA-I00, co-funded by the European Union (FEDER). In addition, NVT receives funding from the Alzheimer’s Disease Data Initiative (ADDI) through the Williams H. Gates Sr. Fellowship Program and the Ajuntament de Barcelona in collaboration with “la Caixa” Foundation through the project 23S06083-001. This work has been conducted within the framework of the PhD program in Biochemistry, Molecular Biology, and Biomedicine at the Autonomous University of Barcelona.

## CONSENT STATEMENT

The ALFA+ study was approved by the independent ethics committee of Parc de Salut Mar Barcelona and has been registered as a clinical trial (identifier: NCT02485730). All study participants provided written informed consent for study participation.

## Supporting information

### List of supplementary tables

**Table S1.** Descriptives of study sample stratified by APOE-e4 status. Legend: 1. Median (Q1, Q3); n (%); 2. Wilcoxon rank sum test; Fisher’s exact test. Mean (SD) follow-up time was 3.45 (0.58) years.

**Table S2.** Associations between LTL and CSF biomarkers at baseline and the change over time.

**Table S3.** Interactions between APOE-e4 status and LTL on the association with CSF biomarkers.

**Table S4.** Stratified analyses by APOE-e4 and AT status on the association between LTL and CSF biomarkers.

**Table S5.** Independence of Aβ status in the associations between LTL and CSF biomarkers in APOE-e4 carriers at baseline.

**Table S6.** Independence of T status in the associations between LTL and CSF biomarkers in APOE-e4 carriers at baseline.

**Table S7.** Interaction of AT status with LTL on the association with CSF biomarkers.

**Table S8.** Independence of CSF AB42/40, ptau181 and NfL in the associations between LTL and AD and aging cortical thickness signatures.

### List of supplementary figures

**Figure S1**. Significant interaction between LTL and APOE-e4 status (Pint = 0.047) on the association with CSF YKL-40 levels at baseline (N = 346). Model was adjusted for age and sex.

**Figure S1.** Significant interaction between LTL and APOE-e4 status (Pint = 0.047) on the association with CSF YKL-40 levels at baseline (N = 346). Model was adjusted for age and sex.

**Figure S2.** Significant interaction between LTL and APOE-e4 status (Pint = 0.036) on the association with the change in CSF sTREM2 levels over time (N = 237). Model was adjusted for age and sex.

**Figure S3.** Significant associations between LTL and log-transformed CSF biomarkers levels at baseline in APOE-e4 carriers (N = 184). Models were adjusted for age and sex.

**Figure S4.** Interaction between LTL and AT status on CSF Aβ42/40 levels at baseline (A-T+ vs. A-T-: Pint = 0.002; A+T-vs. A-T-: P = 0.09) (N = 346). Model was adjusted for age, sex and APOE-e4.

**Figure S5.** Significant associations between LTL and change in (A) CSF α-synuclein and (B) CSF IL-6 levels in the A-T-group. Models were adjusted for age, sex, APOE-e4 status and time difference between the two visits.

**Figure S6.** Significant associations between LTL and baseline levels of (A) CSF NfL, (B) CSF S100B, and (C) CSF YKL-50 at baseline in the A+T-group. Models were adjusted for age, sex, APOE-e4 status.

**Figure S7.** Significant associations between LTL and baseline levels of (A) CSF t-tau, (B) CSF α-synuclein, (C) CSF sTREM2 and change over time of (D) CSF sTREM2 in the A+T+ group. Models were adjusted for age, sex and APOE-e4 status. Model was further adjusted for time difference between visits for the change in CSF biomarkers levels over time.

**Figure S8.** Mediator role of CSF glial and inflammatory biomarkers on the association between leukocyte telomere length and aging and AD cortical thickness signatures. Models were adjusted for age, sex, APOE-e4 status and firmware MRI version. All β are standardized coefficients.

## Abbreviations

Aβ: amyloid-β
AD: Alzheimer’s disease
AT: amyloid β-tau staging
CAIDE: Cardiovascular Risk Factors, Aging, and Incidence of Dementia
CDR: Clinical Dementia Rating
CSF: cerebrospinal fluid
CU: cognitively unimpaired
CV: coefficient of variation
DNA: deoxyribonucleic acid
FDR: False Discovery Rate
GFAP: glial fibrillary acidic protein
IL-6: interleukin 6
LTL: leukocyte telomere length
MCI: mild cognitive impairment
MMSE: Mini-Mental State Examination
MRI: magnetic resonance imaging
NfL: neurofilament light
PET: positron emission tomography
p-tau181: phosphorylated tau181
RT-qPCR: real-time polymerase chain reaction-based assay
sTREM2: soluble triggering receptor expressed on myeloid cells 2
TL: telomere length
t-tau: total tau
YKL-40: chitinase-3-like protein 1

