## Supporting information Figures for "Shorter leukocyte telomere length is associated with distinct CSF biomarker dynamics across early AD stages in at-risk individuals"

### Supporting materials

*Supplementary figures*

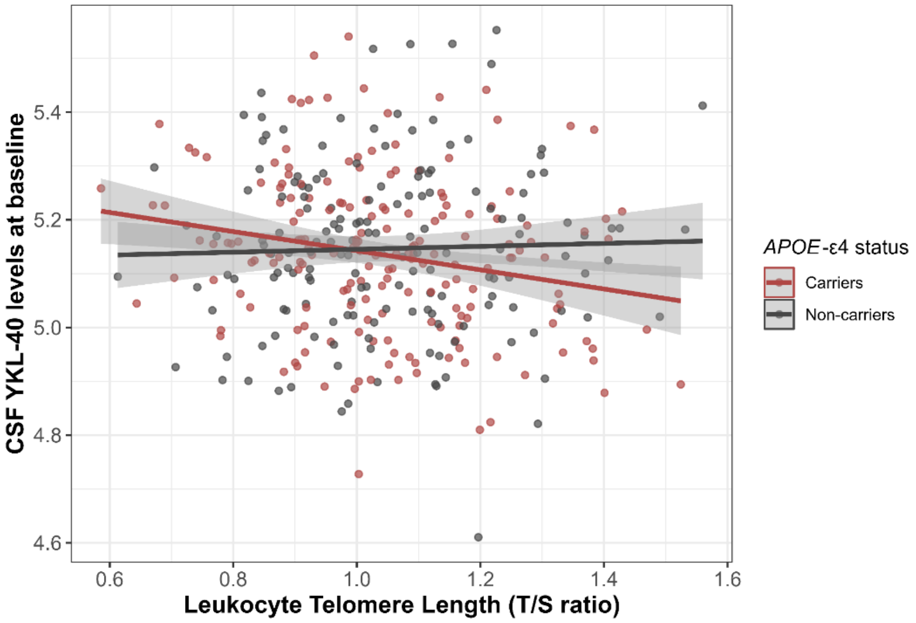


**Figure S1.** Significant interaction between LTL and *APOE*-ε4 status (P*int* = 0.047) on the association with CSF YKL-40 levels at baseline (N = 346). Model was adjusted for age and sex.

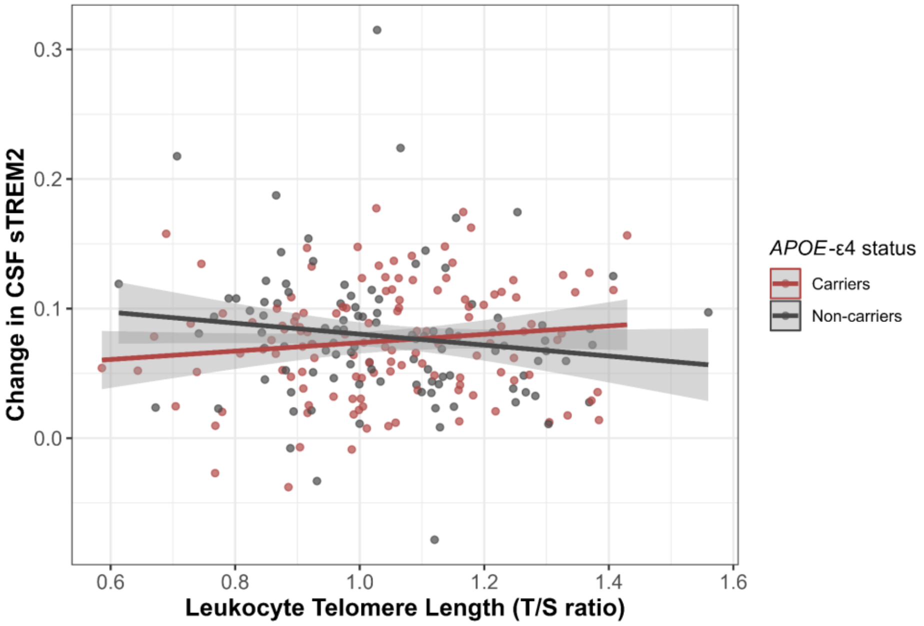


**FigureS2.** Significant interaction between LTL and *APOE*-ε4 status (P*int* = 0.036) on the association with the change in CSF sTREM2 levels over time (N = 237). Model was adjusted for age and sex.

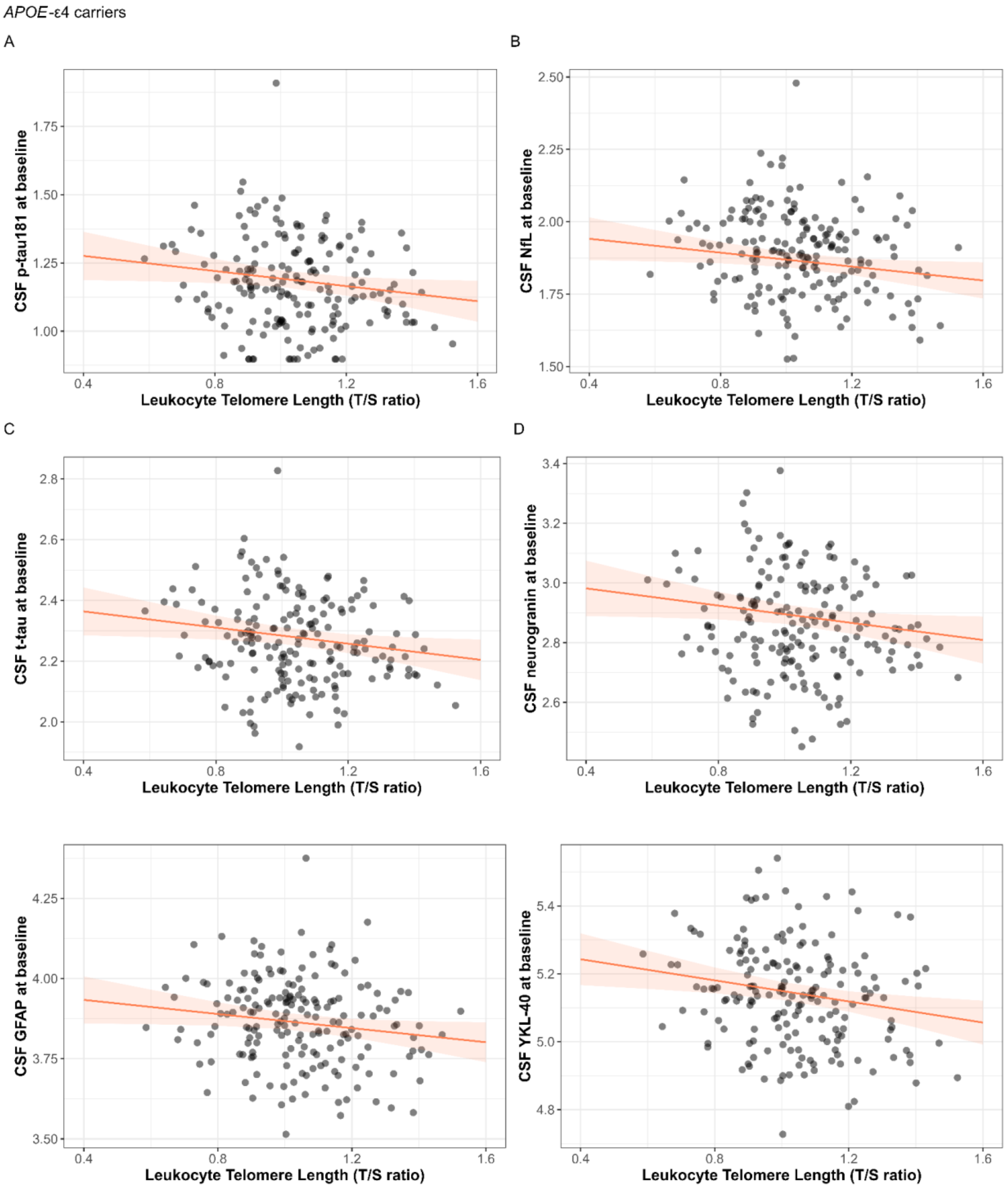


**FigureS3.** Significant associations between LTL and log-transformed CSF biomarkers levels at baseline in *APOE*-ε4 carriers (N = 184). Models were adjusted for age and sex.

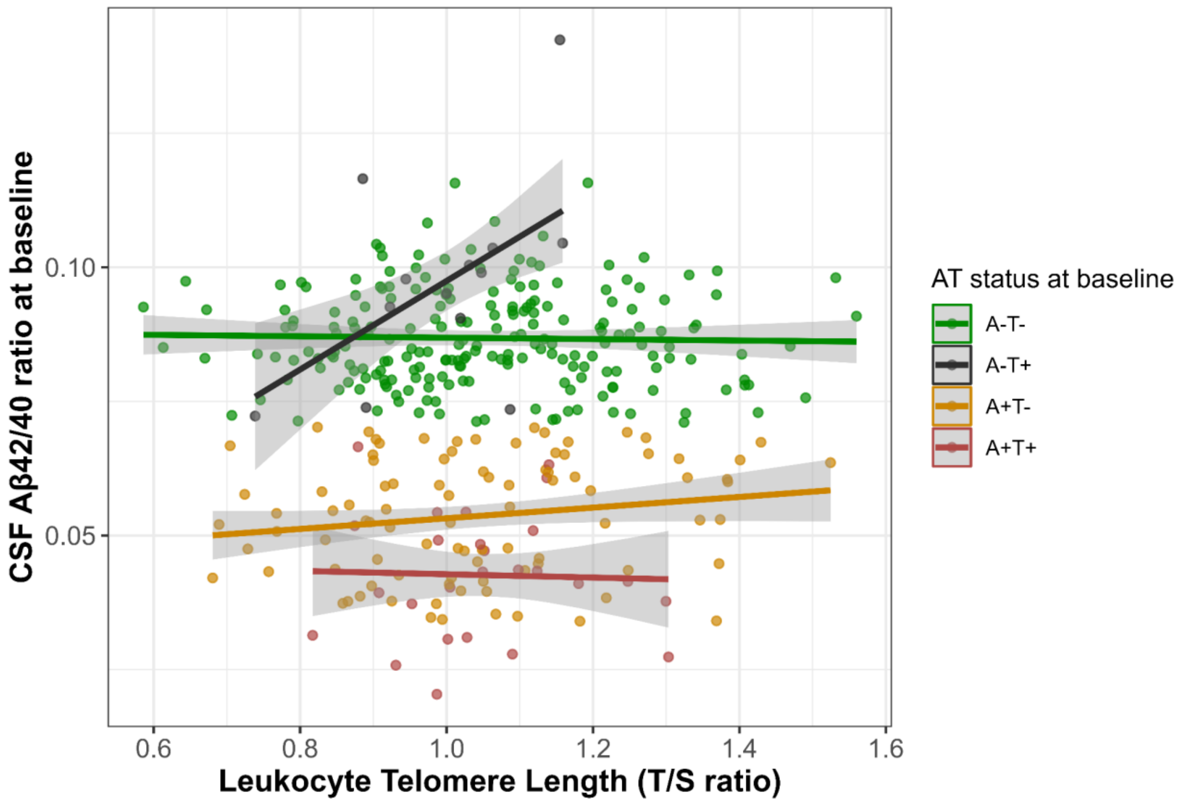


**FigureS4.** Interaction between LTL and AT status on CSF Aβ42/40 levels at baseline (A-T+ vs. A-T-: Pint = 0.002; A+T- vs. A-T-: P = 0.09) (N = 346). Model was adjusted for age, sex and *APOE*-ε4.

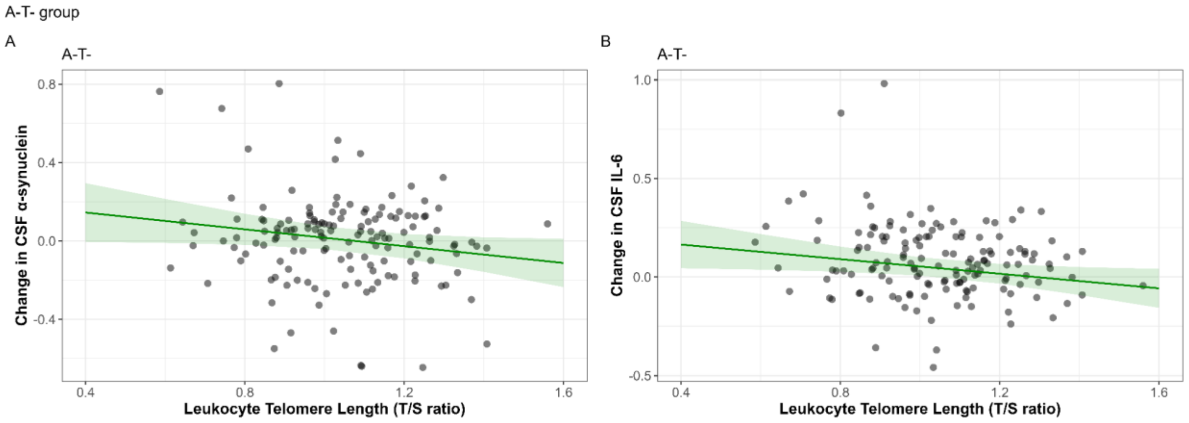


**FigureS5.** Significant associations between LTL and change in (A) CSF α-synuclein and (B) CSF IL-6 levels in the A-T- group. Models were adjusted for age, sex, *APOE*-ε4 status and time difference between the two visits.

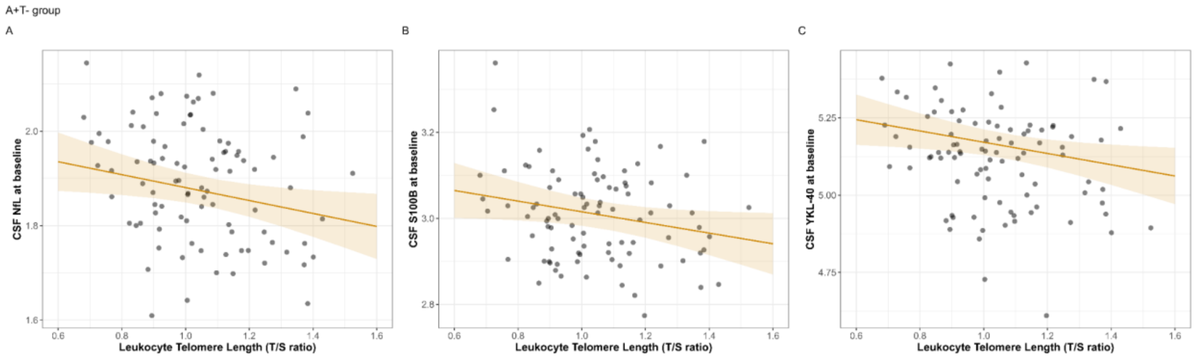


**FigureS6.** Significant associations between LTL and baseline levels of (A) CSF NfL, (B) CSF S100B, and (C) CSF YKL-50 at baseline in the A+T- group. Models were adjusted for age, sex, *APOE*-ε4 status.

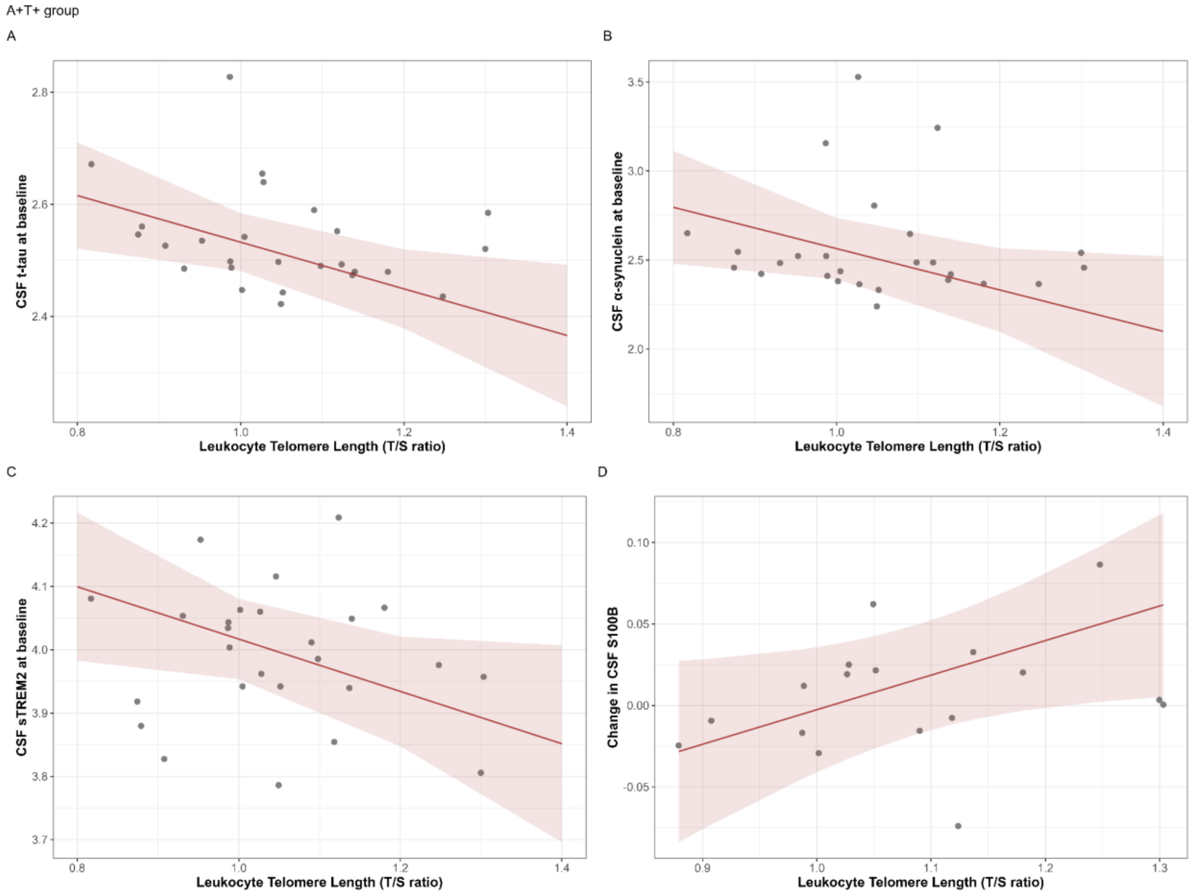


**FigureS7.** Significant associations between LTL and baseline levels of (A) CSF t-tau, (B) CSF α-synuclein, (C) CSF sTREM and change over time of (D) CSF sTREM2 in the A+T+ group. Models were adjusted for age, sex and *APOE*-ε4 status. Model was further adjusted for time difference between visits for the change in CSF biomarkers levels over time.

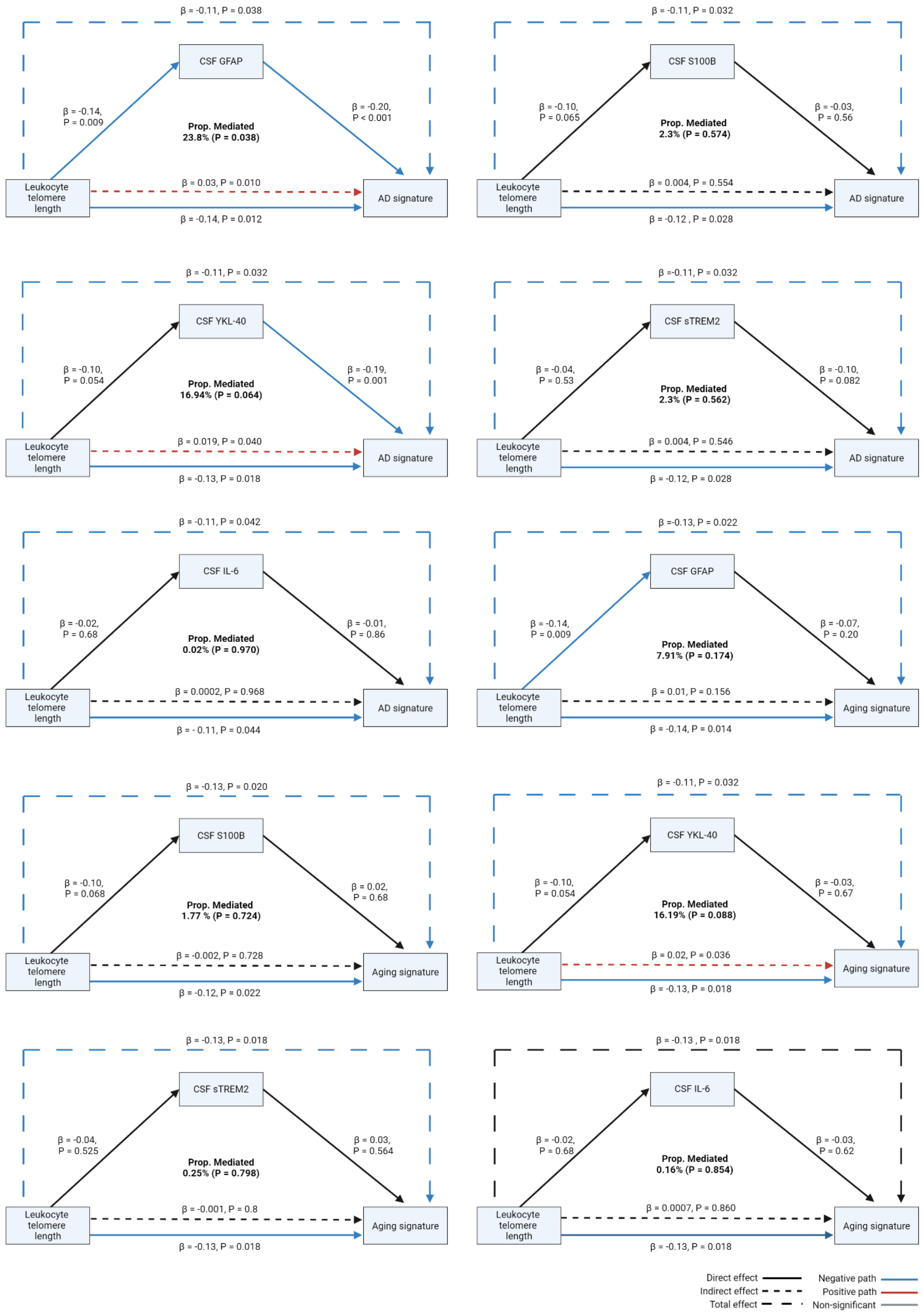


**FigureS8.** Mediator role of CSF glial and inflammatory biomarkers on the association between leukocyte telomere length and aging and AD cortical thickness signatures. Models were adjusted for age, sex, *APOE*-ε4 status and firmware MRI version. All β are standardized coefficients.
